# Influence of sex on disease severity in children with COVID-19 and Multisystem Inflammatory Syndrome in Latin America

**DOI:** 10.1101/2021.02.07.21251212

**Authors:** Martin Brizuela, Jacopo Lenzi, Rolando Ulloa-Gutiérrez, Antúnez-Montes Omar Yassef, Jorge Alberto Rios Aida, Olguita del Aguila, Erick Arteaga-Menchaca, Francisco Campos, Fadia Uribe, Andrea Parra Buitrago, Lina Maria Betancur Londoño, Jessica Gómez-Vargas, Adriana Yock-Corrales, Danilo Buonsenso

**Affiliations:** Pediatric Infectious Disease, Hospital isidoro Iriarte, Quilmes, Buenos Aires, Argentina; Department of Biomedical and Neuromotor Sciences, Alma Mater Studiorum - University of Bologna, Bologna, Italy; Infectious Disease Department. Hospital Nacional de Niños “Dr. Carlos Sáenz Herrera”, CCSS, San José, Costa Rica; Departamento de Docencia e Investigación, Instituto Latinoamericano de Ecografía en Medicina (ILEM)., Ciudad de Mexico, Mexico; CLÍNICA JAS MÉDICA, Lima, Perù; Unidad de Infectología Pediátrica del Hospital Nacional Edgardo Rebagliati Martins-Lima-Perú; Hospital General Regional 200 IMSS, Mexico; Hospital Madre Niño San Bartolome, Lima, Peru; Hospital Pablo Tobon Uribe Medellin, Colombia; Fundacion Neumologica Colombiana,Bogotà, Colombia; Pediatric Emergency Department, Hospital Nacional de Niños “Dr. Carlos Sáenz Herrera”, CCSS, San José, Costa Rica; Department of Woman and Child Health and Public Health, Fondazione Policlinico Universitario A. Gemelli, Rome, Italy; Dipartimento di Scienze Biotecnologiche di Base, Cliniche Intensivologiche e Perioperatorie, Università Cattolica del Sacro Cuore, Rome, Italy; Global Health Research Institute, Istituto di Igiene, Università Cattolica del Sacro Cuore, Roma, Italia

**Keywords:** COVID-19, MIS-C, Sex, Gender

## Abstract

Data from adult studies how that COVID-19 is more severe in men than women. However, no data are available for the pediatric population. For this reason, we performed this study aiming to understand if sex influenced disease severity and outcomes in a large cohort of latin-american children with COVID-19 and Multisystem Inflammatory Syndrome (MIS-C). We found that a higher percentage of male children developed MIS-C (8.9% vs 5% in females) and died (1.2% and 0.4% in females), although on multivariate adjusted analyses the only statistically significant difference was found in need of hospitalization, with females less frequently admitted compared with boys (25.6% vs 35.4%). This data are preliminary and need further independent studies to better assess the role of sex.

## INTRODUCTION

More than a year after the description of the first cases of COVID-19 in China, several aspects of this pandemic are still unclear. Among these, there is the different clinical impact of COVID-19 in women and men. Early data from China showed that men were more frequently infected by COVID-19 than women, and men with underlying diseases (diabetes, hypertension and cardiovascular disease) developed severe condition with an increased mortality rate (1). Similar findings were reported from Italy and, subsequently, confirmed almost in all countries.

Several hypothesis have been postulated to support these differences, although definite conclusions have not reached yet. Disparities in in sex-specific disease outcomes may be due to sex-specific steroids and activity of X-linked genes which modulate the innate and adaptive immune response to virus infection and influence the immune response (2). The sex-related pre-existing comorbidities, such as hypertension, cardiovascular disease and diabetes, can play a role, since they were associated with severe outcomes and are more frequent in men (3). Also, hormonal and genetic factors can affect ACE2 expression, the receptor of SARS-CoV-2 (4-6), microRNAs expressions and transcription (7), and Vitamin D3 activity (8).

However, if such a differences between sexes is present in the pediatric population has never been analyzed yet. Major pediatric papers mainly focused age as a risk factors of disease severity but, to our knowledge, no sub-analyses assessing sex have been described (9-15). Since sex-hormones or specific habits (alcohol, type-2 diabetes, cardiovascular disease) are less pronounced in the pediatric population, in particular in infants and young children, such an analyses can provide indirect evidence of influence of hormones on COVID-19. Therefore, we performed this study aiming to understand if sex influenced disease severity and outcomes in a large cohort of latin-american children with COVID-19 and Multisystem Inflammatory Syndrome (MIS-C).

## MATERIALS AND METHODS

### Study Design and Participants

This study is part of an ongoing independent project assessing COVID-19 and MIS-C in Latin American children, already presented elsewhere (12) and with a previous published paper describing an initial group of 409 children with confirmed COVID-19 (14). For the current study, we performed a sub-analyses of a previously used dataset (15) aiming to assess the influence of sex on disease severity. The remaining variables are those previously described and included age, gender, symptoms, imaging, underlying medical conditions, need for hospital and NICU/PICU admission, respiratory and cardiovascular support, other viral co-infections, drugs used to treat COVID-19, development of MIS-C and type of organ involvement, and outcome.

SARS-CoV-2 infection was defined as a positive PCR test on nasopharyngeal swab.

MIS-C was defined according to the CDC criteria: An individual aged <21 years (we only included if younger than 18 years) presenting with i) fever, ii)laboratory evidence of inflammation, and iii) evidence of clinically severe illness requiring hospitalization, with multisystem (>2) organ involvement (cardiac, renal, respiratory, hematologic, gastrointestinal, dermatologic or neurological); and iiii) no alternative plausible diagnoses; and iiiii) positive for current or recent SARS-CoV-2 infection by RT-PCR, serology, or antigen test; or exposure to a suspected or confirmed COVID-19 case within the 4 weeks prior to the onset of symptoms.

The study was reviewed and approved by the CoviD in sOuth aMerIcaN children—study GrOup core group and approved by the Ethics Committee of the coordinating center and by each participating center (Mexico: COMINVETICA-30072020-CEI0100120160207; Colombia: PE-CEI-FT-06; Peru: No. 42-IETSI-ESSALUD-2020; Costa Rica: CEC-HNN-243-2020). The study was conducted in accordance with the Declaration of Helsinki and its amendments. No personal or identifiable data were collected during the conduct of this study.

### Statistical analysis

Summary statistics were presented as counts and percentages. The association between female sex and COVID-19 clinical outcomes was preliminarily evaluated with crude odds ratios (ORs) and 95% confidence intervals (CIs). Confounding adjustment was performed using a propensity score approach based on inverse-probability weighting. More specifically, we considered the following covariates potentially related with the outcomes and unbalanced across males and females: age, pre-existing medical conditions, immunosuppressants at the time of diagnosis, primary or secondary immunodeficiency, chemotherapy over the last 6 months, pyrexia (≥38.0/≥100.4 °C/°F), days between symptom onset and diagnosis, administration of systemic corticosteroids, intravenous immunoglobulin therapy, lower respiratory tract infection, and MIS-C diagnosis. Clustered standard errors were used to account for the multicenter design of the study. All data were analyzed using the Stata 15 software (StataCorp. 2017. *Stata Statistical Software: Release 15*. College Station, TX: StataCorp LLC). The significance level was set at 5% and all tests were 2-sided.

## RESULTS

The characteristics of the 990 patients included in the study, overall and by sex, are summarized in Table 1. Among the 484 females, 39 (8.1%) had chest X-ray abnormalities, 124 (25.6%) were admitted to the hospital, 20 (4.1%) were admitted to ICUs, 49 (10.1%) received respiratory support, and 2 (0.4%) died. 24 (5%) were diagnosed with MIS-C. Among 506 males, 53 (10.5%) had chest X-ray abnormalities, 179 (35.4%) were admitted to the hospital, 27 (5.3%) were admitted to ICUs, 69 (13.6%) received respiratory support, and 6 (1.2%) died. 45 (8.9%) were diagnosed with MIS-C. Results of crude and adjusted analysis showing the association between sex and clinical outcomes are shown in Table 2. Following adjustment obtained with a weighting approach based on propensity scores, the only significant outcome was access to the hospital: girls were admitted less frequently than boys (OR = 0.82, P <0.001).

**Table 1.**
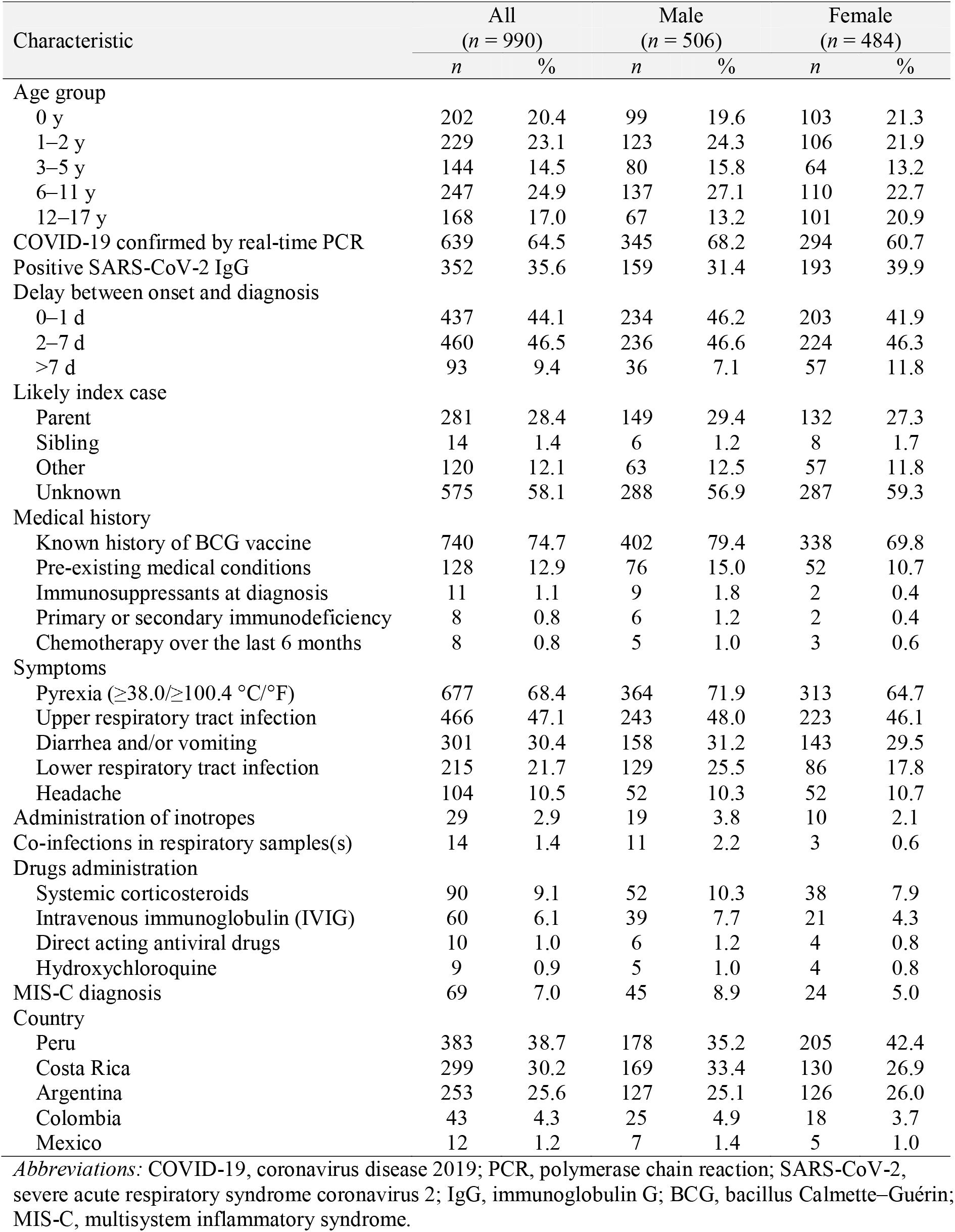
Characteristics of the study sample, overall and by sex.

**Table 2.** Crude and adjusted association between female sex and COVID-19 clinical outcomes, expressed as odds ratios (ORs) and 95% confidence intervals (CI).

| Outcome | *n* (%) | Crude analysis |  |  | Adjusted analysis* |  |  |
| --- | --- | --- | --- | --- | --- | --- | --- |
|  |  | OR | 95% CI | *P*-value | OR | 95% CI | *P*-value |
| Chest X-ray abnormalities† | 92 (9.3) | 0.75 | 0.58–0.96 | 0.022 | 1.09 | 0.86–1.36 | 0.480 |
| Hospital admission | 303 (30.6) | 0.63 | 0.55–0.72 | <0.001 | 0.82 | 0.78–0.86 | <0.001 |
| ICU admission | 47 (4.7) | 0.76 | 0.64–0.92 | 0.004 | 1.15 | 0.94–1.40 | 0.173 |
| Respiratory support‡ | 118 (11.9) | 0.71 | 0.58–0.88 | 0.001 | 1.08 | 0.87–1.34 | 0.487 |
| Death | 8 (0.8) | 0.34 | 0.07–1.65 | 0.183 | 0.43 | 0.09–2.10 | 0.297 |

\*Confounding adjustment obtained with a weighting approach based on propensity scores.
†Pneumonia and/or acute respiratory distress syndrome.
‡Oxygen support, mechanical ventilation and/or continuous positive airway pressure therapy.
Abbreviation: ICU, intensive care unit.

## DISCUSSION

In this study, we aimed to assess the impact of sex on disease severity in a large cohort of Latin American children with COVID-19 and MIS-C. We found that a higher percentage of male children developed MIS-C (8.9% vs 5% in females) and died (1.2% and 0.4% in females), although on multivariate adjusted analyses the only statistically significant difference was found in need of hospitalization, with females less frequently admitted compared with boys (25.6% vs 35.4%). Taken all together, these preliminary data highlight that females can experience a milder disease compared with boys, as suggested in adults. In other reports (6), the females were more affected by hyposmia or anosmia and gustatory dysfunction compared with males; unfortunately, we did not assess this parameters in this study. In our cohort, children had no comorbidities such as hypertension, obesity or type-2 diabetes, therefore this behavioral factors, traditionally more frequent in males, should have not contributed in the described differences. Also, considering that sex hormones may have less impact in children, therefore different microRNAs expressions due to sex-hormones should not be involved as well.

An explanation in of disparity in sex-specific disease outcomes can be found in the activity of X-linked genes, as previously suggested (6), which modulate the innate and adaptive immune response to virus infection and influence the immune response. Also, the male predominance in the COVID-19 pandemic could partially be explained by sex-specific expressions of TMPRSS2 (6).

The activity and expression of the human angiotensin-converting enzyme 2 (ACE2), the functional receptor for the severe acute respiratory syndrome coronavirus (SARS-CoV), can also explain some differences. Age-related differences have been already reported in a review (16). Also, hormonal and genetic factors could lead to ACE2 over-expression in female sex. Gagliardi et al (6) reported that infection with SARS-CoV induces ACE2 down-regulation through binding of the viral Spike protein to ACE2, thus reducing ACE2 expression in the lung and igniting acute respiratory failure (17).

Vitamin D has also received attention in the immune pathogenesis of COVID-19 (17) and Pagano et al hypothesiezed that the synergy between Vitamin D3 and estrogen could affect the sex differences in the outcome of patients with COVID-19 (18, 19). However, no rigorous data are yet available on this issues and, in our series, Vitamin D levels have not been reported.

Sex may play an important, major role in severity and length of persisting symptoms after the first diagnosis of acute COVID-19. In adults, long covid has been described and it seems more frequent in women (20-21). In children, international experts highlighted the possibility of this disease in childhood as well and recent report seemed to confirm that long covid may affect children as well (22, 23). Considering the autoimmunity has been suggested as a potential pathogenic trigger and the higher frequency of autoimmune disorders in children, the role of sex in long covid, including children, is a priority research topic for the following months.

Our study has limitation to address. First, it was not initially developed to specifically address how sex could influence disease severity, therefore the variables included as well the study power was not tailored to this topic. Also, blood tests, hormones and Vitamin D were not tested and therefore not analyzed. Last, there is no a control group of adult patients from the same area to compare sex differences in different age groups. However, this study is the first one specifically assessing the role of sex in children with COVID-19 and MIS-C.

In conclusion, we found that a slightly more severe course of COVID-19 and MIS-C in boys compared with women in our cohort of Latin American children. This data are preliminary and need further independent studies to better assess the role of sex. In light of the growing evidence of long covid in children, it is important to begin including sex as an important potential variable of severity or symptoms persistence in children with COVID-19.

## Data Availability

available upon request to the corresponding author

## Ethic approval

approved by each institution (codes provided in methods)

## Availability of data and material

available upon request

